# Analysis of Augmentation Techniques for Spine X-Ray Images

**DOI:** 10.64898/2026.04.15.26350121

**Authors:** Elakiya Sivakumar, Anjana Anand

## Abstract

Computer vision and deep learning techniques, including convolutional neural networks (CNNs) and transformers, have increased the performance of medical image classification systems. However, training deep learning models using medical images is a challenging task that necessitates a substantial amount of annotated data. In this paper, we implement data augmentation strategies to tackle dataset imbalance in the VinDr-SpineXR dataset, which has a lower number of spine abnormality X-ray images compared to normal spine X-ray images. Geometric transformations and synthetic image generation using Generative Adversarial Networks are explored and applied to the abnormal classes of the dataset, and classifier performance is validated using VGG-16 and InceptionNet to identify the most effective augmentation technique. Additionally, we introduce a hybrid augmentation technique that addresses class imbalance, reduces computational overhead relative to a GAN-only approach, and achieves ∼99% validation accuracy with both classifiers across all three case studies.

## 1. Introduction

Medical image analysis is a well-established computer vision domain encompassing tasks such as disease classification, lesion segmentation, and anatomical landmark detection, with direct clinical applications in screening, diagnosis, monitoring, and treatment planning. The growing volume and complexity of radiological data have driven sustained interest in computer-assisted diagnostics (CAD), in which deep learning models automate feature extraction and inference directly from raw images. Computer vision methods have demonstrated strong performance across diverse medical image classification tasks, continually expanding the range of viable deep learning approaches [1,2].

There are various dataset-related challenges in using deep learning to automate the medical image classification process. Due to the extensive effort involved in gathering and classifying data, assembling large datasets can be a formidable task involving manual work. In medical image analysis, limited datasets are a particularly common problem. The rarity of disorders, patient privacy, the demand of medical professionals for labelling, and the cost and manual effort required to create medical image databases all impose practical limits on achievable dataset scale [2,3]. Dataset bias also arises when the training distribution fails to reflect the diversity of real-world clinical presentations, differences in scanner hardware, acquisition protocol, and patient demographics can introduce systematic discrepancies between training and deployment conditions [2,3].

Deep learning models do not generalise well to new data when trained on restricted datasets, classification methods require large volumes of data to overcome overfitting and class generalisation difficulties. For large-scale clinical applications, models must be trained with diverse, representative datasets. When a disorder type constitutes only a small fraction of the overall dataset, the data are said to be imbalanced. Medical imaging datasets frequently have this problem, making disorder classification extremely challenging: a standard classifier tends to be biased toward the majority class and overlook minority pathologies which are precisely the cases of greatest clinical significance —2 resulting in incorrect patient diagnoses [2]. The issue is compounded by the large file sizes of X-ray, MRI, and CT volumes, which constrain large-scale dataset curation and distribution.

To address these medical imaging dataset challenges, data augmentation is a viable solution. Data augmentation is a set of strategies for producing additional training samples from existing data in order to artificially increase the amount of data available. Image data augmentation transforms training images into label-preserving modified versions, raising the training sample size without requiring additional real-world data collection. This is especially important when annotated data are scarce and acquiring new examples is expensive and time-consuming, as is the case in medical imaging [4].

Degenerative spinal conditions place a significant burden on healthcare systems worldwide, and accurate radiographic diagnosis is critical for effective clinical management [5]. The VinDr-SpineXR dataset [6] consists of substantially more normal images than abnormal spine disorder images, presenting a clear class imbalance problem; spinal disorders represent a significant and growing clinical burden [7]. This study analyses how data augmentation can address this issue, implementing and comparing basic affine-transformation augmentation, synthetic GAN-based augmentation, and a novel hybrid strategy that combines the best-performing techniques of both.

### 1.1 Study objectives

The aim of this study is to analyze different data augmentation methods (basic, synthetic, and hybrid methods) on the VinDr spine dataset and compare the classifier model performance (multi-class classification) before and after image dataset augmentation.

- To obtain images from the VinDr-SpineXR dataset and perform data preprocessing.
- To identify different data augmentation techniques (basic augmentation and GAN-based image generation) to perform on the dataset.
- To implement the different data augmentation techniques on the abnormal classes of VinDr-SpineXR.
- To study the best performing augmentation methodologies and implement a novel “hybrid” augmentation methodology that combines basic augmentation and GAN-based image generation strategies.
- To build multi-class classifier models for performance validation.
- To analyze the performance of the classifiers before and after data augmentation is implemented and observe the classifier network performances for each case.

## 2. Dataset Details

This study uses the VinDr-SpineXR dataset [5], a large-scale annotated collection of 10,466 spine radiographs drawn from 5,000 patient investigations. Plain radiography is the primary clinical tool for detecting and monitoring spinal disorders [8]: it delivers clear visualisation of bony structures at low cost and broad availability, making it well-suited to population-scale dataset construction. Each image in the dataset was manually annotated by a qualified radiologist across 13 disorder categories, and the data were collected at Hanoi Medical University Hospital under an approved institutional review protocol. Figure 1 shows representative images from the dataset.

**Figure 1.**
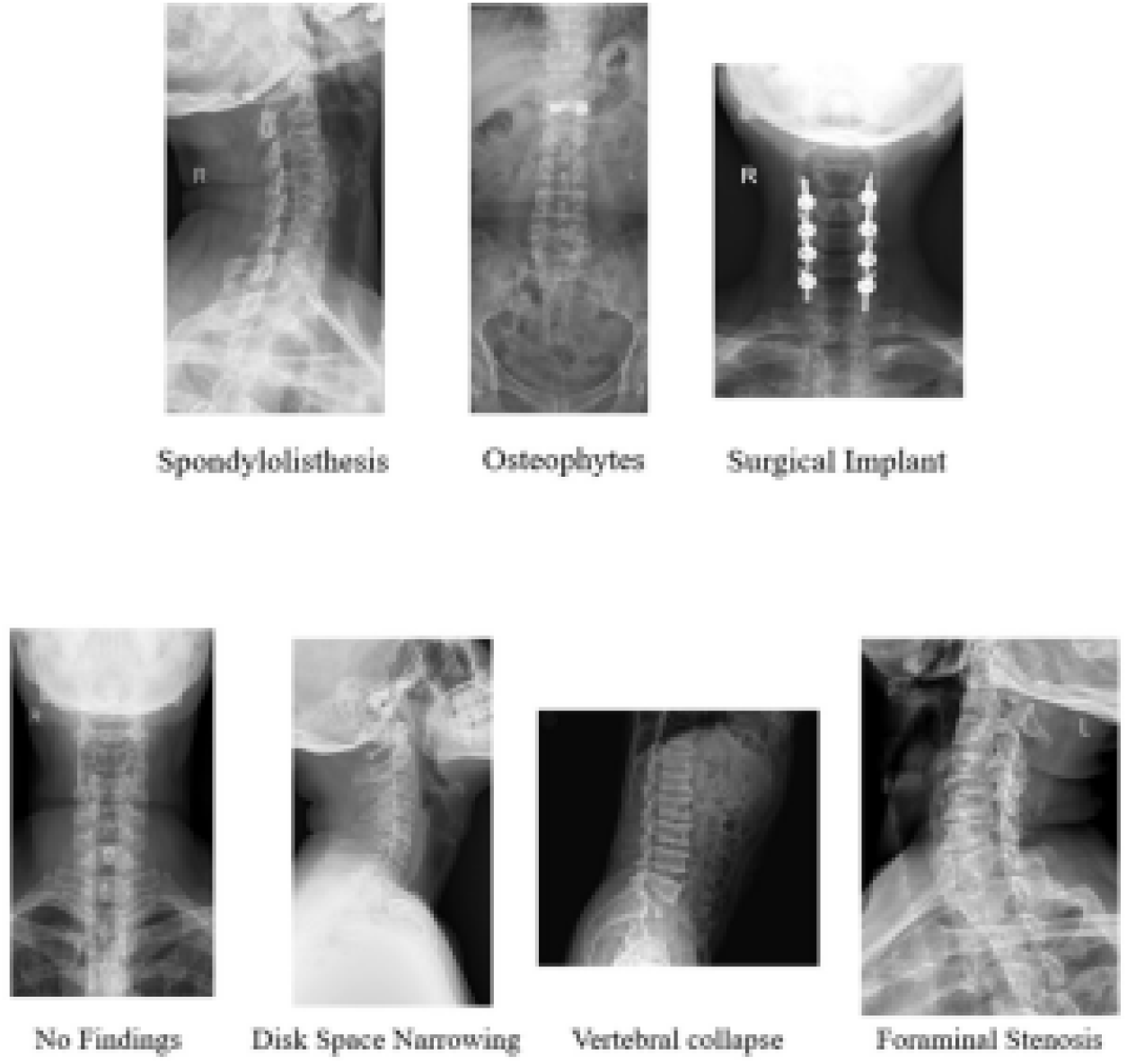
Images from the VinDr-SpineXR dataset.

The dataset was published in August 2021 and was selected for this study on account of its breadth of disorder categories and the pronounced class imbalance it exhibits. As shown in Figure 2, abnormal class images are substantially fewer than No Findings images (healthy spine images).

**Figure 2.**
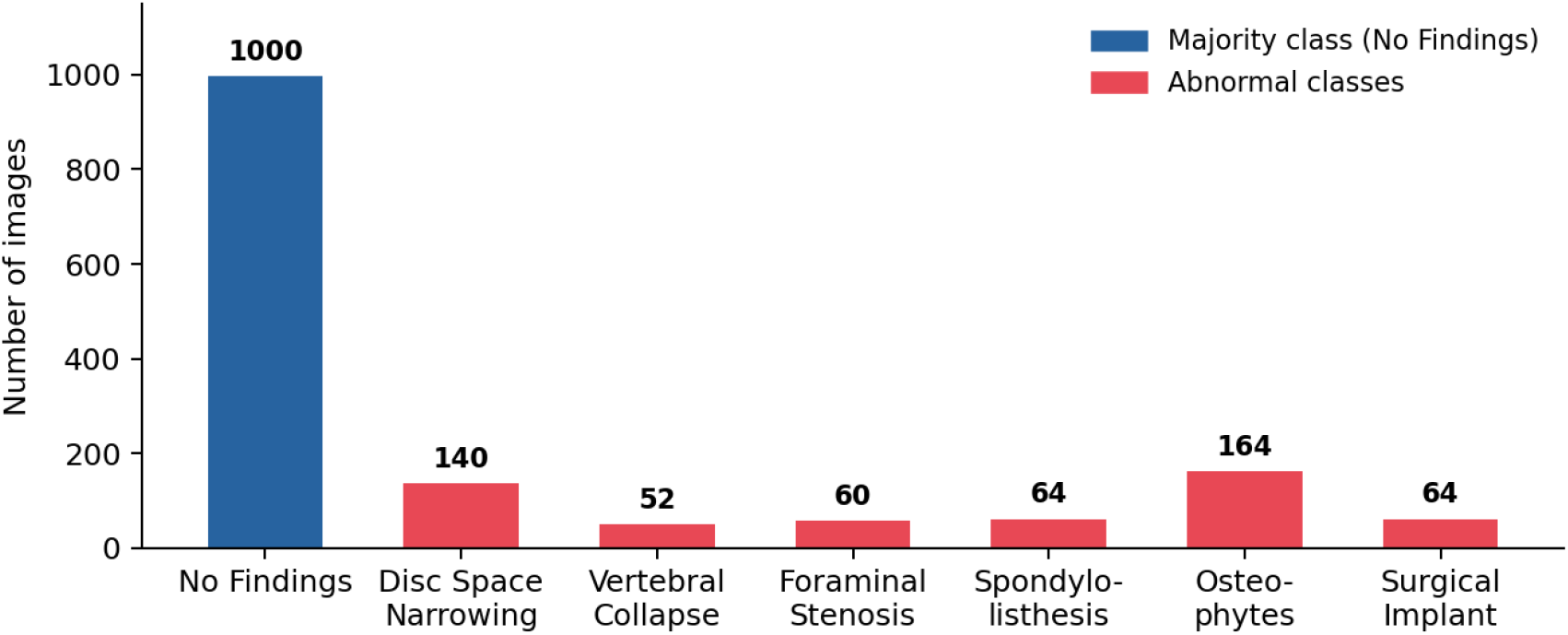
Original class distribution of the VinDr-SpineXR study subset. The No Findings class (n=1,000) substantially outnumbers every abnormal class (range: 52–164), motivating augmentation of the minority classes.

**Figure 3.**
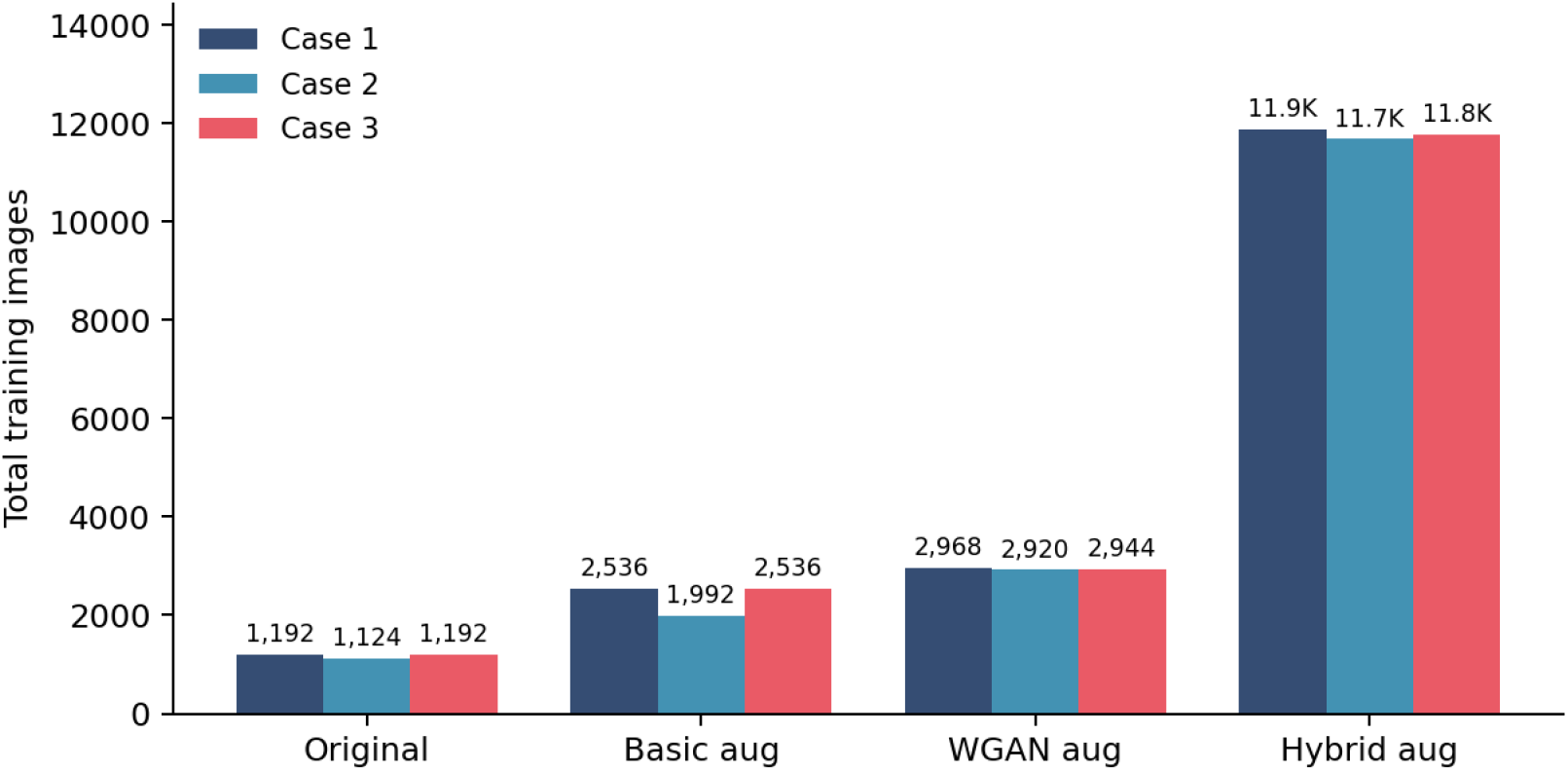
Total training image count at each augmentation stage for Cases 1, 2 and 3. Hybrid augmentation expands the dataset approximately 10× relative to the original dataset, resolving the class imbalance across all cases.

## 3. Literature Survey

Medical image classification is a well-established computer vision task, and deep learning methods, particularly CNNs and transformers [9], have achieved strong performance across diverse anatomical regions and imaging modalities. Data augmentation has matured into a highly pertinent research area, with a growing body of systematic reviews and empirical benchmarks published through the 2020s [2,3,4].

### 3.1 Basic Image Augmentation

Geometric and affine transformation-based augmentation — rotation, flipping, cropping, zooming, and shearing — constitutes the foundational approach to training set expansion in computer vision. Shorten and Khoshgoftaar [2] provide a comprehensive taxonomy of such techniques, and Chlap et al. [4] systematically reviewed their application to medical imaging datasets across CT, MRI, and histopathological modalities. A consistent finding is that augmentation benefit depends strongly on strategy selection rather than volume alone: Hussain et al. [10] demonstrated that Gaussian noise addition degrades mammography classification accuracy, while selective rotation and flip combinations have yielded substantial accuracy improvements on CT classification tasks. This sensitivity of performance to augmentation choice directly motivates the per-strategy empirical evaluation carried out in the present study.

### 3.2 Synthetic Image Augmentation Using GANs

Generative Adversarial Networks [11], and in particular Deep Convolutional GANs (DCGAN [12]) and Wasserstein GANs (WGAN [13]), have made GAN-based synthesis practical for medical imaging by addressing training instability and mode collapse. Frid-Adar et al. [10] demonstrated that DCGAN-synthesised liver lesion images improve classification sensitivity from 78.6% to 85.7%, outperforming classical augmentation alone. The gradient-penalty WGAN variant [13] provides further training stability. Combining GAN-generated images with classical geometric transforms has emerged as an effective hybrid strategy: Sajjad et al. [14] showed that applying rotation and flipping to DCGAN-generated Alzheimer’s PET images improved accuracy from 65% to 72%. Systematic reviews confirm that DCGAN and conditional GAN architectures remain the most widely adopted approaches in medical augmentation pipelines [3], with Vrettos et al. [15] specifically identifying spinal radiography as an underexplored domain for GAN-based augmentation, the gap the present study addresses. Denoising diffusion models represent an emerging alternative paradigm for medical image synthesis [16]; the present study focuses on GAN-based approaches, which were the established generative framework at the time this work was conducted.

The present study draws on these established methods such as selective geometric augmentation, WGAN-based synthesis, and a combined hybrid pipeline to address class imbalance in the VinDr-SpineXR spinal radiograph dataset.

## 4. Methodology

### 4.1 Dataset Preparation

The VinDr-SpineXR dataset [5] contains the spine X-ray images in DICOM format. For easier handling of the images, the DICOM images were converted into JPEG format for compatibility with the Python image processing pipeline; visual inspection confirmed that compression artefacts were not perceptible at the resolutions used. A CSV file mapping image IDs to class labels was also prepared. The VinDr-SpineXR dataset is split into three different case studies for the implementation of augmentation strategies across different cases to understand which technique works best for all cases. The classes in each case and the number of images in each category are outlined in Table 1.

**Table 1.** Different cases employed for the study.

| CASE 1 |  | CASE 2 |  | CASE 3 |  |
| --- | --- | --- | --- | --- | --- |
| CATEGORY | IMAGES | CATEGORY | IMAGES | CATEGORY | IMAGES |
| NO FINDINGS | 1000 | NO FINDINGS | 1000 | NO FINDINGS | 1000 |
| DISC SPACE NARROWING | 140 | FORAMINAL STENOSIS | 60 | OSTEOPHYTES | 164 |
| VERTEBRAL COLLAPSE | 52 | SPONDYLOLISTHESIS | 64 | SURGICAL IMPLANT | 64 |

### 4.2 Classifier Networks

The networks were implemented in the Python language in a Conda–Jupyter environment. Keras with a TensorFlow backend was employed in building and training the networks. The networks were designed to perform multiclass classification between the no-finding and the two abnormal classes respectively.

VGG-16 and InceptionNet were implemented because of their established performance in computer vision [17, 18]. Although primarily validated on natural image benchmarks, both architectures have been widely adopted in medical image classification tasks and provide a well-understood baseline for augmentation comparison studies.

For both networks, baseline hyperparameters were set after analysing early stopping trends with raw data. The number of epochs was set to 15; the learning rate was set at 0.0001 to maximize accuracy, minimize loss and training time. For consistency in comparison these baselines are followed throughout the study.

### 4.3 Training

Each case was first run through the classifier networks (VGG-16 and InceptionNet) without any image augmentation, and baseline accuracies were recorded for the unaugmented dataset. This is referred to as vanilla training and serves as the reference against which augmentation improvements are measured.

The dataset was then augmented using basic affine transformation techniques applied only to the abnormal classes, directly targeting the class imbalance. All seven basic strategies were first applied in combination; each strategy was also applied individually to identify which techniques contribute positively to classifier performance.

Next, synthetic augmentation was performed on the abnormal classes using GANs for all three cases. The GAN-augmented datasets were then passed through the classifier networks and performance was recorded. GAN implementation details are provided in §4.6.

Finally, a novel hybrid augmentation strategy was introduced, combining data warping (affine transforms) with oversampling (GAN synthesis). The best-performing geometric strategies identified by the individual augmentation analysis (§5.3) were applied to the GAN-augmented dataset to maximise the number of training images. The hybrid-augmented datasets for all three cases were passed through the classifier networks and performance was analysed. Figure 4 provides an overview of the full experimental workflow.

**Figure 4.**
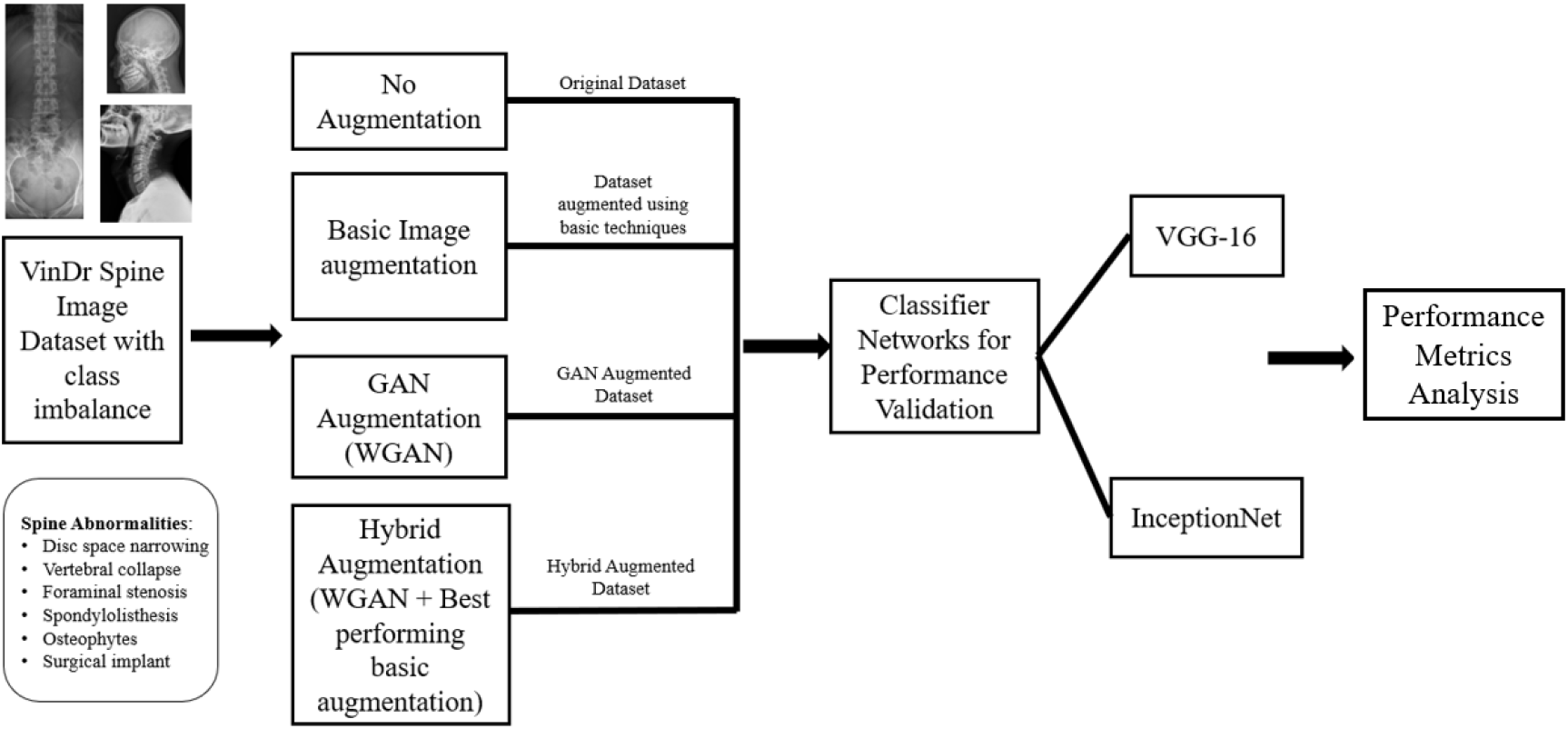
Study workflow

### 4.4 Affine Augmentation

An affine transformation is a function that preserves the structure of images while mapping an object from one affine space to another. An affine transformation retains lines or distance ratios while altering the image’s orientation, size, or position. A transformation matrix is used for image alteration through affine transformation methods. To increase the dataset size of the abnormal classes, seven geometric transformations were applied: rotation (90° and 270° for Case 1; 45° and 270° for Cases 2 and 3), horizontal flip, vertical flip, cropping, zooming, and 30° shearing.

### 4.5 Individual Augmentation Analysis

Each of the seven geometric augmentation strategies was also applied independently to each case dataset in Python, in order to isolate each technique’s individual contribution to classifier performance. Each augmented image was saved as a JPEG file and passed separately to the classifier networks.

### 4.6 Generative Adversarial Networks

A Generative Adversarial Network consists of two competing networks known as the Generator and Discriminator. The input to the GAN is the real samples. The generator’s goal is to produce images similar to the real-world samples while the discriminator’s goal is to differentiate between real and fake samples.

For each epoch, both the generator and discriminator are trained. The discriminator is trained first and then the generator. As the generator improves with training, the discriminator performance gets worse because the discriminator cannot easily tell the difference between real and fake. If the generator succeeds perfectly, then the discriminator has 50% accuracy (like flipping a coin). The input to the generator is a latent vector — a point in a high-dimensional noise space sampled from a Gaussian distribution at runtime — which the generator network maps to a synthetic image. The latent vector size used in this study was 120.

Generators and discriminators each have their losses. During training the losses for both networks are minimised through adversarial competition until the generator produces images the discriminator can no longer reliably distinguish from real samples, at which point the discriminator approaches 50% accuracy. To compute this loss, traditionally min–max / binary cross-entropy loss is applied.

#### 4.6.1 DCGANs

Due to its prominence in image data augmentation, Deep Convolutional GAN (DCGAN) was implemented first. The discriminator and generator of DCGANs use convolutional and convolutional-transpose blocks, respectively.

The convolutional transpose layer, dropout, and Leaky ReLU activation function make up the generator’s upsampling block. In the discriminator, the downsampling block consists of convolutional, batch normalization, and Leaky ReLU activation function. The stride was set to (2, 2) and kernel size to (5, 5). Binary cross-entropy loss was implemented and the network was trained for 200 epochs. The latent dimensions were set to 120, and the generator produces 64×64 output images.

#### 4.6.2 Wasserstein GANs (WGAN)

WGAN was introduced in 2017 [12] with the basic architecture of a DCGAN and utilizing the Wasserstein loss. The W-loss or the earth mover’s distance (EMD) is a measure of the distance between two probability distributions, in this case the generated image and the real images. WGANs are able to overcome stability issues, mode collapse, and minimize the need for hyperparameter tuning. This is achieved through implementing W-loss, which provides the GAN more stability during training by relaxing the importance of the discriminator which provides binary output, and defines a “critic” based on the Earth Mover’s Distance. However, the distances are not minimized below a threshold so as to reduce copies of the reference image and large clashes between the discriminator loss and the W-loss.

In the implementations, latent dimensions of size 120 and images of size 256×256 were used. The algorithm used for implementing WGAN using the PyTorch package is shown in Figure 6. Representative outputs at selected training epochs are shown in Figure 7. The generator and discriminator loss curves, along with image quality metric progression across training epochs, are shown in Figure 8.

**Figure 6.**
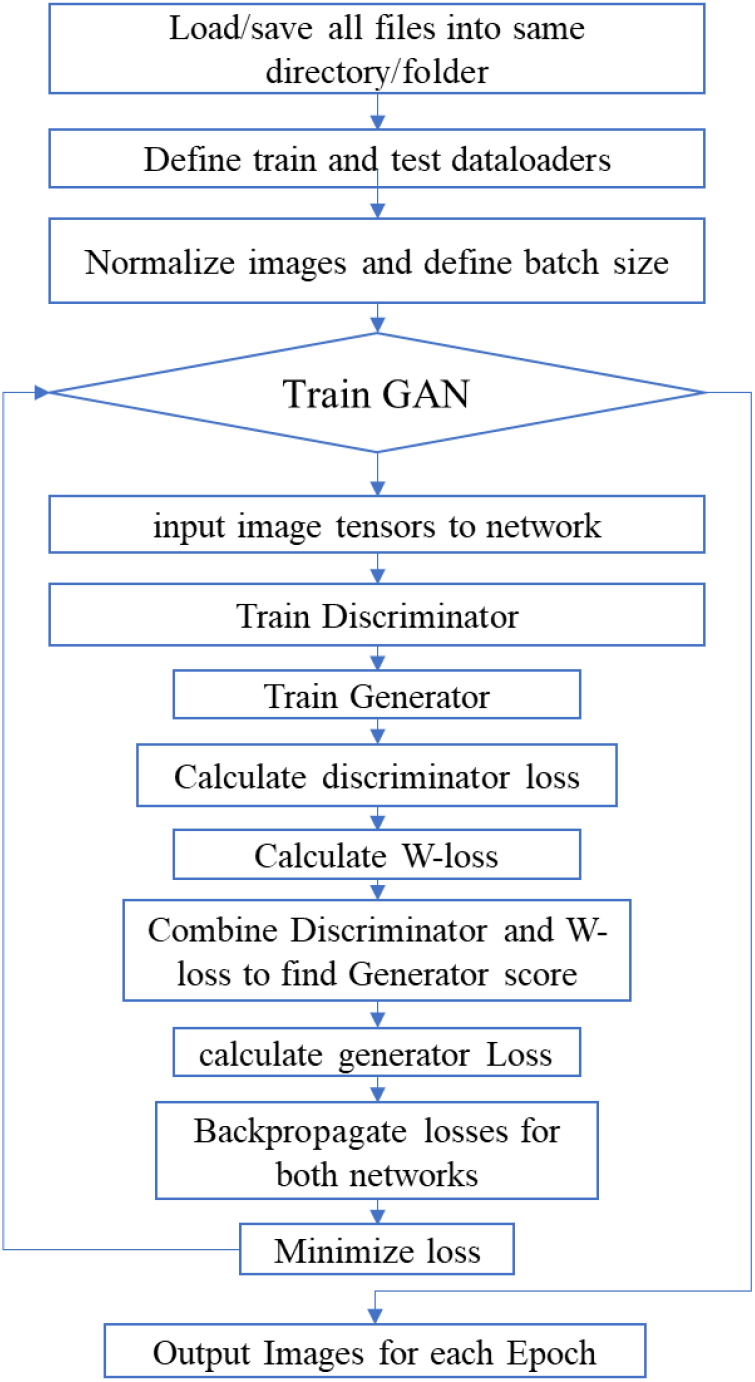
WGAN training algorithm implemented in PyTorch. Latent dimension 120, image size 256×256, Wasserstein loss with weight clipping.

**Figure 7.**
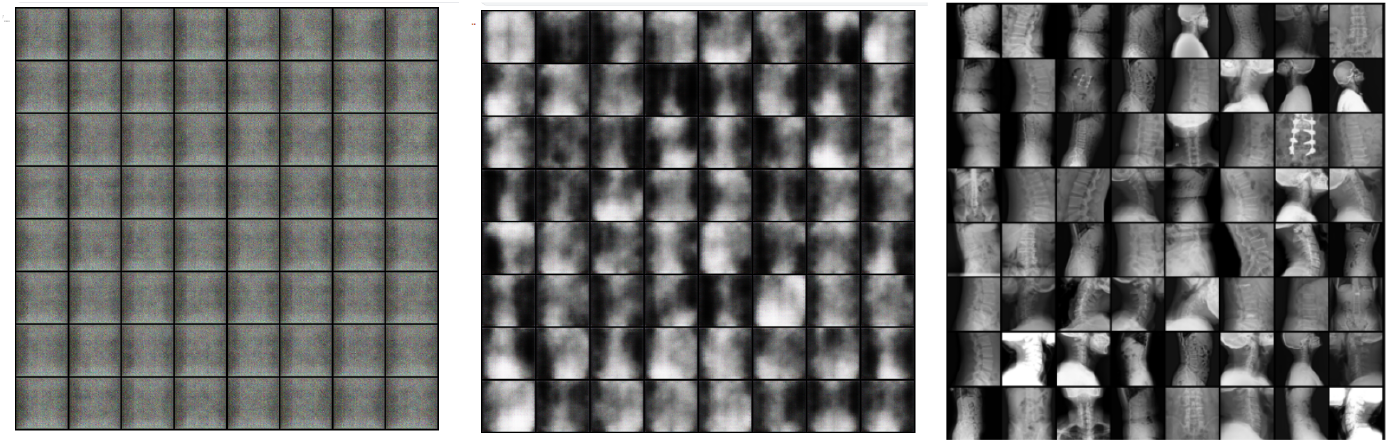
WGAN synthetic image quality progression for Case 1. Left to right: Epoch 1, Epoch 100, Epoch 200. Images become progressively more realistic; quality degradation was observed in extended training beyond approximately the 300th epoch.

**Figure 8.**
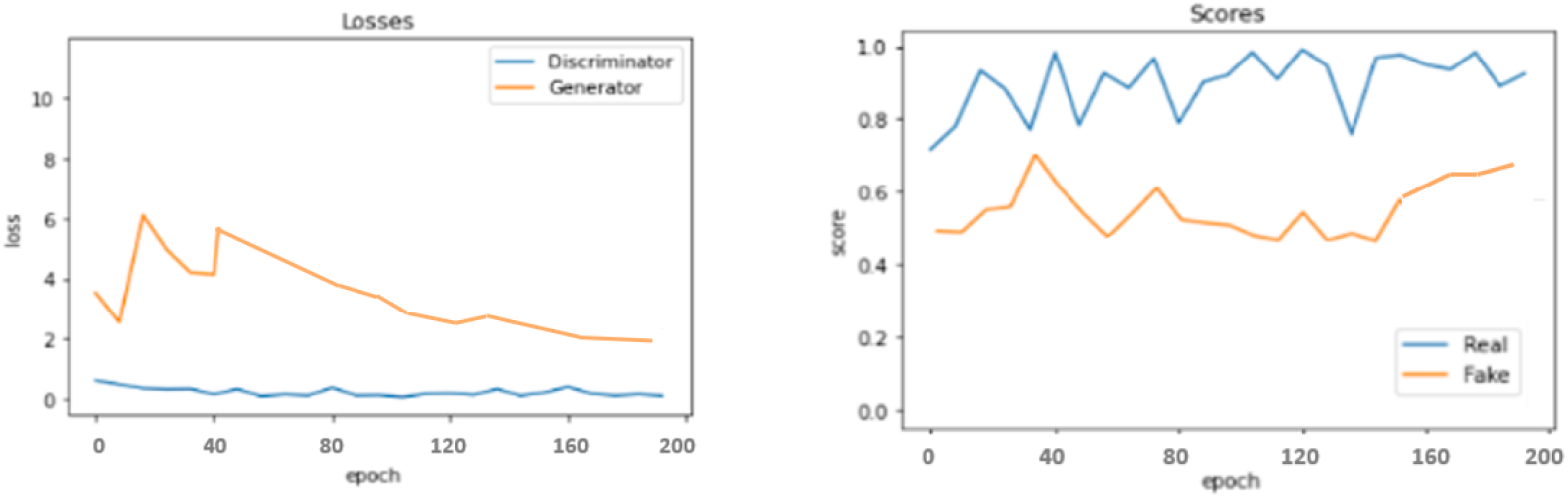
WGAN generator and discriminator loss curves during training (left) and real vs. fake score progression (right). The W-loss serves as a training signal and a proxy for generator stability.

## 5. Results

Results are reported across four augmentation conditions-vanilla training (no augmentation), basic geometric augmentation, WGAN-based synthetic augmentation, and hybrid augmentation was evaluated with both VGG-16 and InceptionNet classifiers across all three case studies. Section 5.1 reports unaugmented baseline classifier performance. Section 5.2 presents combined basic augmentation classifier performance. Section 5.3 reports individual augmentation technique results. Section 5.4 presents WGAN-only classifier performance. Section 5.5 reports hybrid augmentation results.

An overview of the dataset size at each augmentation stage is provided in Figure 3. WGAN augmentation expanded the abnormal class counts by 148%, 159%, and 139% for Cases 1, 2, and 3 respectively, resolving the class imbalance that constrained vanilla training. The following subsections detail results for each augmentation strategy in turn.

### 5.1 Unaugmented Baseline

Each case was first evaluated without any augmentation to establish the reference performance against which augmentation improvements are measured. Table 2 reports classifier accuracy for VGG-16 and InceptionNet across all three cases under vanilla (no augmentation) training.

**Table 2.** Unaugmented baseline classifier performance.

| Case | No. of images | VGG16 Train | VGG16 Val | Inception Train | Inception Val |
| --- | --- | --- | --- | --- | --- |
| Case 1 (Disc space narrowing + Vertebral collapse) | 1,192 | 80.63% | 70.00% | 79.66% | 78.28% |
| Case 2 (Foraminal stenosis + Spondylolisthesis) | 1,124 | 85.27% | 86.59% | 84.39% | 85.66% |
| Case 3 (Osteophytes + Surgical implant) | 1,228 | 80.26% | 81.21% | 81.27% | 81.43% |

The pronounced class imbalance in Case 1 (140 and 52 abnormal images against 1,000 normal images) is reflected in the lower VGG-16 validation accuracy of 70%, demonstrating that a standard classifier trained on this imbalanced distribution is biased toward the majority class.

### 5.2 Combined Basic Augmentation

Applying all seven geometric strategies to the abnormal classes together produced 2,536, 1,992, and 2,824 training images for Cases 1, 2, and 3 respectively. Table 3 reports classifier accuracy for VGG-16 and InceptionNet across all three cases under combined basic augmentation. Combined basic augmentation yielded up to ∼24 percentage points improvement in VGG-16 validation accuracy for Case 1 (from the 70% baseline) and ∼4–5 percentage points for Cases 2 and 3; InceptionNet showed ∼5–6 percentage point improvements across all cases.

**Table 3.** Combined basic augmentation classifier performance.

| Case | No. of images | VGG16 Train | VGG16 Val | Inception Train | Inception Val |
| --- | --- | --- | --- | --- | --- |
| Case 1 (Disc space narrowing + Vertebral collapse) | 2,536 | 93.32% | 94.07% | 84.28% | 83.09% |
| Case 2 (Foraminal stenosis + Spondylolisthesis) | 1,992 | 89.69% | 90.39% | 88.17% | 89.59% |
| Case 3 (Osteophytes + Surgical implant) | 2,824 | 85.81% | 85.56% | 87.28% | 88.09% |

### 5.3 Individual Augmentation Techniques

Each augmentation technique was applied independently across all three cases and evaluated with both classifier networks to identify which strategy contributes most to classifier performance.

#### Case 1

Results for Case 1 are shown in Table 4.

**Table 4.**
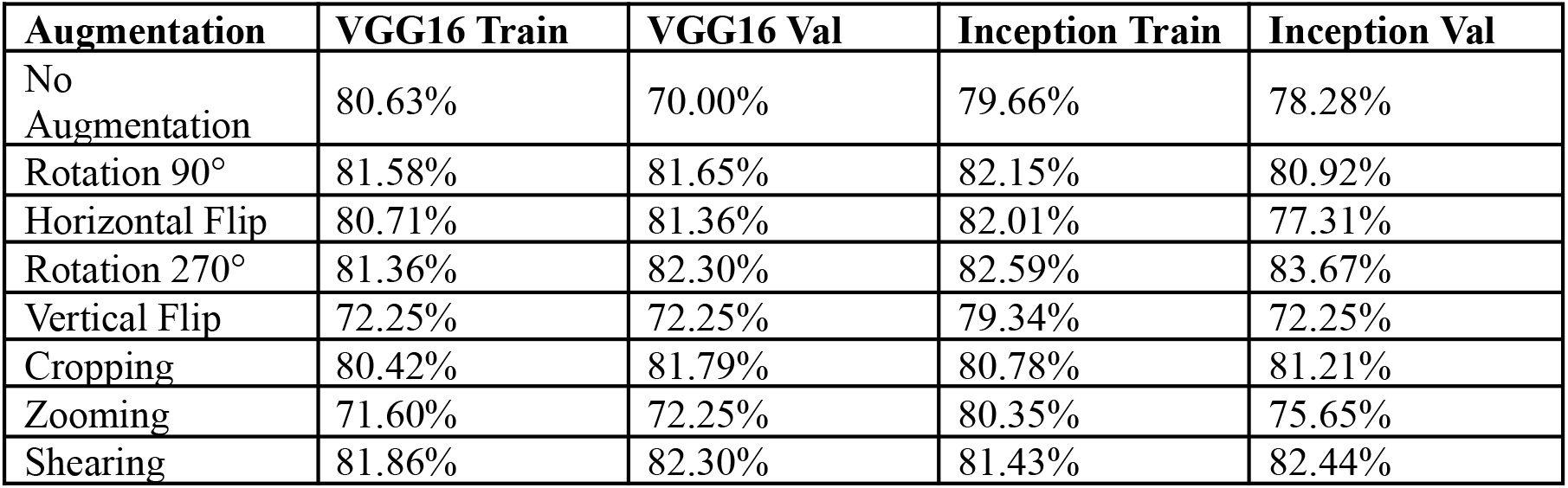
Individual augmentation technique results, Case 1.

| Augmentation | VGG16 Train | VGG16 Val | Inception Train | Inception Val |
| --- | --- | --- | --- | --- |
| No Augmentation | 80.63% | 70.00% | 79.66% | 78.28% |
| Rotation 90° | 81.58% | 81.65% | 82.15% | 80.92% |
| Horizontal Flip | 80.71% | 81.36% | 82.01% | 77.31% |
| Rotation 270° | 81.36% | 82.30% | 82.59% | 83.67% |
| Vertical Flip | 72.25% | 72.25% | 79.34% | 72.25% |
| Cropping | 80.42% | 81.79% | 80.78% | 81.21% |
| Zooming | 71.60% | 72.25% | 80.35% | 75.65% |
| Shearing | 81.86% | 82.30% | 81.43% | 82.44% |

#### Case 2

Results for Case 2 are shown in Table 5.

**Table 5.** Individual augmentation technique results, Case 2.

| Augmentation | VGG16 Train | VGG16 Val | Inception Train | Inception Val |
| --- | --- | --- | --- | --- |
| No Augmentation | 85.27% | 86.59% | 84.39% | 85.66% |
| Rotation 45° | 85.80% | 87.95% | 85.36% | 88.36% |
| Horizontal Flip | 86.28% | 87.26% | 85.40% | 87.76% |
| Rotation 270° | 86.80% | 88.31% | 85.28% | 88.21% |
| Vertical Flip | 82.54% | 82.14% | 83.01% | 83.81% |
| Cropping | 85.28% | 86.87% | 84.36% | 85.99% |
| Zooming | 82.80% | 82.31% | 83.28% | 83.21% |
| Shearing | 86.47% | 88.20% | 85.94% | 86.48% |

#### Case 3

Results for Case 3 are shown in Table 6.

**Table 6.**
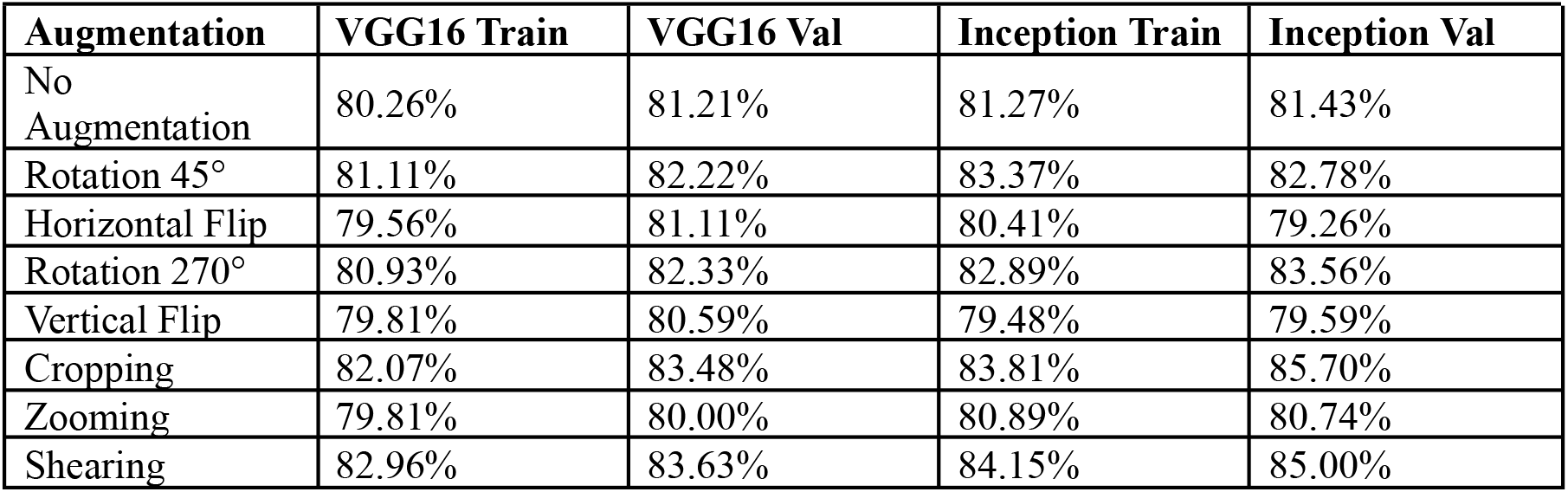
Individual augmentation technique results, Case 3.

| Augmentation | VGG16 Train | VGG16 Val | Inception Train | Inception Val |
| --- | --- | --- | --- | --- |
| No Augmentation | 80.26% | 81.21% | 81.27% | 81.43% |
| Rotation 45° | 81.11% | 82.22% | 83.37% | 82.78% |
| Horizontal Flip | 79.56% | 81.11% | 80.41% | 79.26% |
| Rotation 270° | 80.93% | 82.33% | 82.89% | 83.56% |
| Vertical Flip | 79.81% | 80.59% | 79.48% | 79.59% |
| Cropping | 82.07% | 83.48% | 83.81% | 85.70% |
| Zooming | 79.81% | 80.00% | 80.89% | 80.74% |
| Shearing | 82.96% | 83.63% | 84.15% | 85.00% |

Applying these individual basic strategies across all three cases with both classifiers, three strategies consistently result in better network performances across cases: Rotation 270° and Shearing in all cases, with Rotation 90° the best first rotation for Case 1 and Rotation 45° for Cases 2 and 3. This also validated the hypothesis that combining the best performing strategies together results in better network performance than combining the worst performing strategies together.

### 5.4 WGAN Augmentation

DCGAN was trained independently on each abnormal class subset prior to WGAN. Despite hyperparameter adjustments, DCGAN was unable to learn generalizable features for the VinDr-SpineXR data: generated images exhibited incoherent structure throughout training and showed no recognisable pathological anatomy. DCGAN-generated images were therefore excluded from classifier evaluation.

WGAN was trained independently for each abnormal class subset, using images from the final epochs before quality degradation. In an extended training experiment of up to 500 epochs, over-sharpened outputs were observed beyond approximately the 300th epoch due to stationary critics. Images were therefore selected from epochs prior to this threshold. Table 7 reports WGAN-only classifier performance across all three cases.

**Table 7.** WGAN augmentation classifier performance.

| Case | No. of images | VGG16 Train | VGG16 Val | Inception Train | Inception Val |
| --- | --- | --- | --- | --- | --- |
| Case 1 (Disc space narrowing + | 2,968 | 97.02% | 97.07% | 92.06% | 91.85% |
| Vertebral collapse) |  |  |  |  |  |
| Case 2<br>(Foraminal stenosis + Spondylolisthesis) | 2,920 | 94.28% | 95.09% | 94.62% | 93.19% |
| Case 3<br>(Osteophytes + Surgical implant) | 2,944 | 91.67% | 90.38% | 92.02% | 91.78% |

WGAN augmentation substantially outperformed combined basic augmentation across all cases. The dataset was augmented by approximately 148%, 159%, and 139% for Cases 1, 2, and 3 respectively, providing VGG-16 validation accuracies of 97.07%, 95.09%, and 90.38%, and InceptionNet validation accuracies of 91.85%, 93.19%, and 91.78%.

### 5.5 Hybrid Augmentation

#### Case 1, Hybrid: Rotation 90° + Rotation 270° + Shear

Total images: original 1192 → after WGAN 2968 → hybrid 11,872. Training and validation accuracy curves are shown in Figure 9a. Table 8 summarises classifier performance.

**Table 8.** Hybrid augmentation classifier performance, Case 1.

| Model | No. of images | VGG16 Train | VGG16 Val | Inception Train | Inception Val |
| --- | --- | --- | --- | --- | --- |
| Without Augmentation | 1192 | 80.63% | 70% | 79.66% | 78.28% |
| Basic Augmentation | 2536 | 93.32% | 94.07% | 84.28% | 83.09% |
| WGAN | 2968 | 97.02% | 97.07% | 92.06% | 91.85% |
| **Hybrid Augmentation** | **11872** | **99.19%** | **99.31%** | **98.43%** | **99.13%** |

**Figure 9a.**
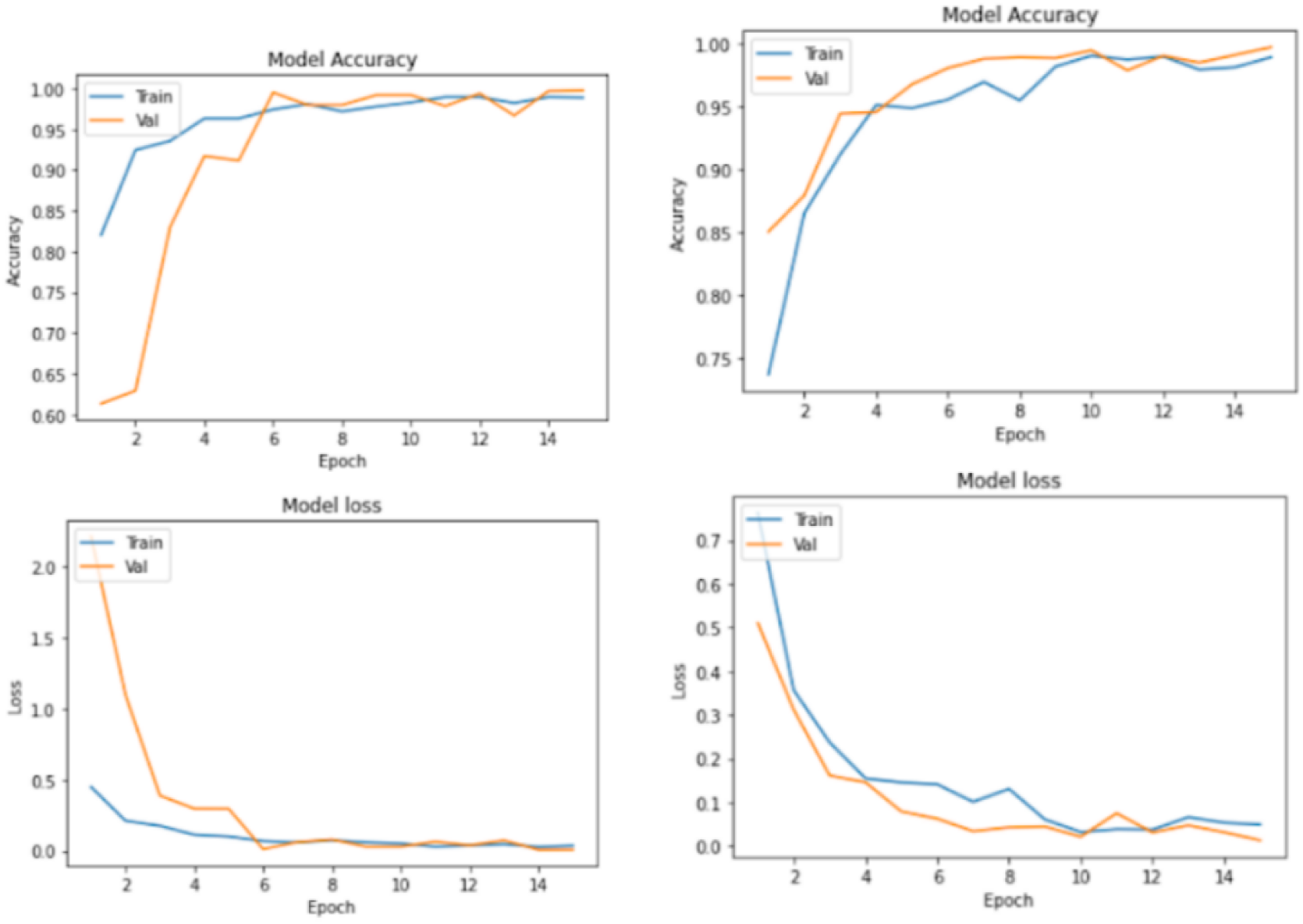
Hybrid augmentation training and validation accuracy curves for Case 1, VGG-16 (left) and InceptionNet (right). Both networks reach ∼99% validation accuracy.

#### Case 2, Hybrid: Rotation 45° + Rotation 270° + Shear

Total images: original 1124 → after WGAN 2920 → hybrid 11,680. Training and validation accuracy curves are shown in Figure 9b. Table 9 summarises classifier performance.

**Figure 9b.**
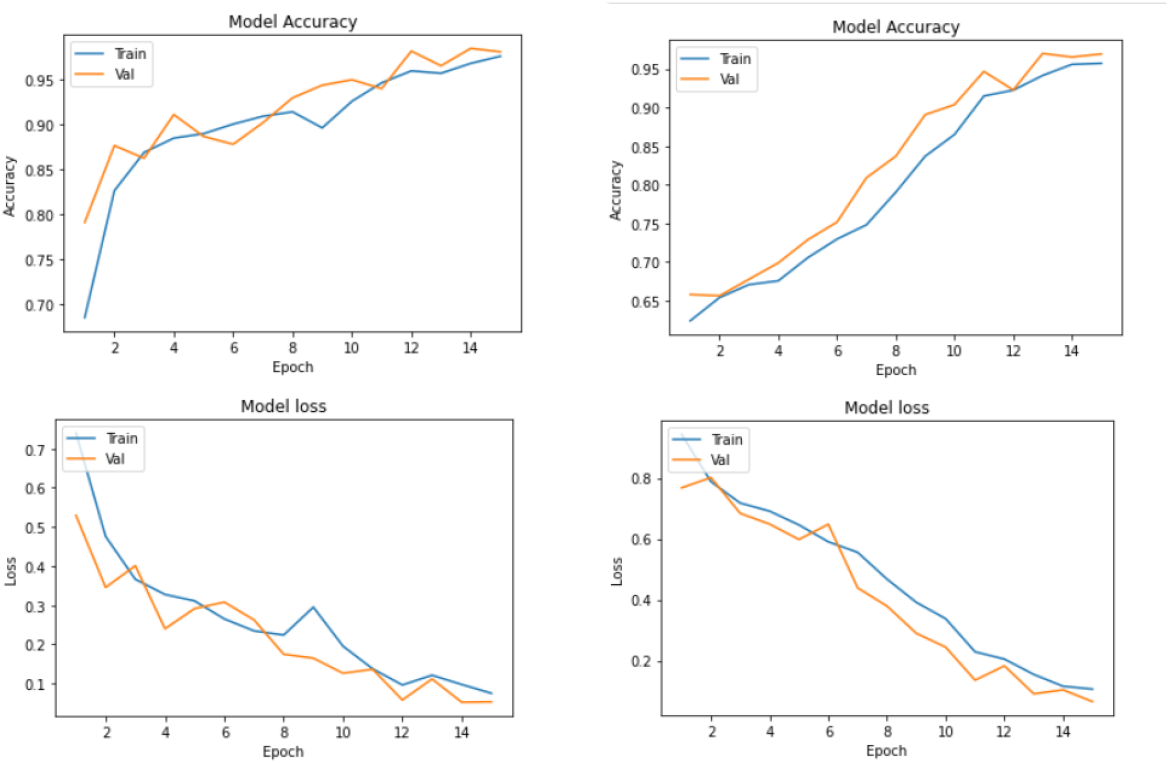
Hybrid augmentation training and validation accuracy curves for Case 2, VGG-16 (left) and InceptionNet (right).

**Table 9.**
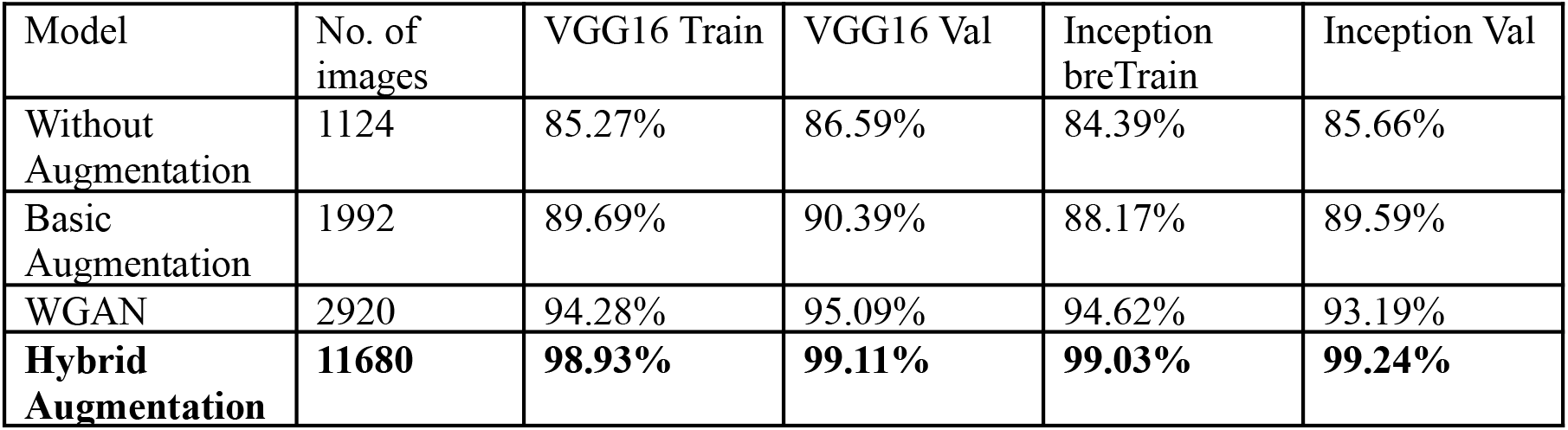
Hybrid augmentation classifier performance, Case 2.

#### Case 3, Hybrid: Rotation 45° + Rotation 270° + Shear

Total images: original 1,228 → after WGAN 2944 → hybrid 11,776. Training and validation accuracy curves are shown in Figure 9c. Table 10 summarises classifier performance.

**Figure 9c.**
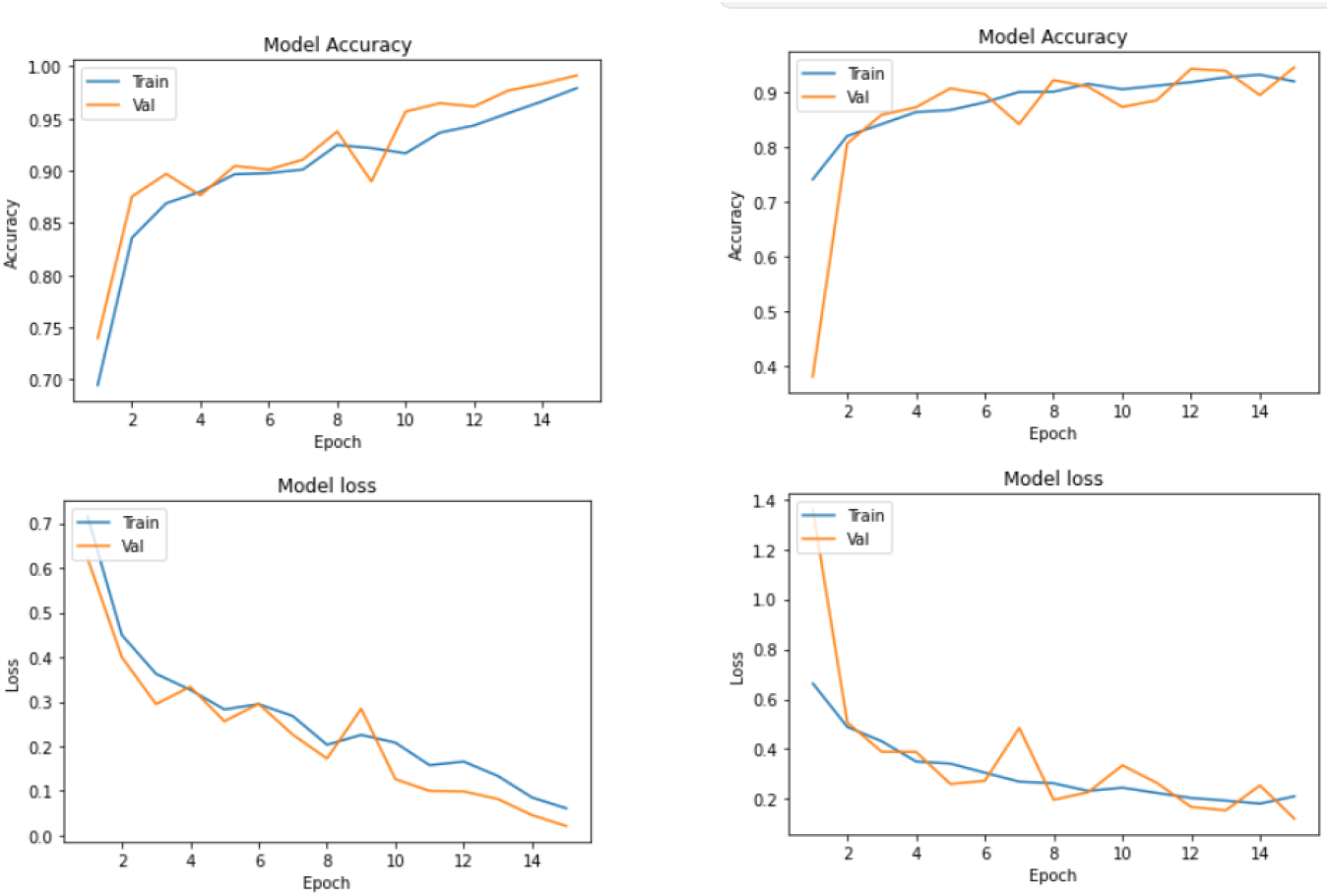
Hybrid augmentation training and validation accuracy curves for Case 3, VGG-16 (left) and InceptionNet (right).

**Table 10.** Hybrid augmentation classifier performance, Case 3.

| Model | No. of images | VGG16 Train | VGG16 Val | Inception Train | Inception Val |
| --- | --- | --- | --- | --- | --- |
| Without Augmentation | 1228 | 80.26% | 81.21% | 81.27% | 81.43% |
| Basic Augmentation | 2824 | 85.81% | 85.56% | 87.28% | 88.09% |
| WGAN | 2944 | 91.67% | 90.38% | 92.02% | 91.78% |
| **Hybrid Augmentation** | **11776** | **98.63%** | **99.21%** | **98.83%** | **99.34%** |

With the novel hybrid augmentation method, abnormal spine disorder images were first generated using WGAN to overcome dataset imbalance. Next, the three best-performing geometric strategies were applied to the GAN-augmented dataset: Rotation 270° and Shearing (common to all cases), plus Rotation 90° for Case 1 and Rotation 45° for Cases 2 and 3. This method generated ∼11K images per case (∼35K in total across all three cases) and yielded network accuracies of ∼99% for both classifiers.

## 6. Discussion

This study systematically compared basic geometric, GAN-based, and hybrid augmentation strategies for addressing class imbalance in a multi-class spinal x-ray classification task. Evaluation was conducted across three case studies using VGG-16 and InceptionNet as validation classifiers, allowing assessment of augmentation effects across architectures of differing depth.

VGG-16 and InceptionNet were selected as validation classifiers on the basis of their established use in traditional classification problems and their contrasting architectural profiles, shallow-and-wide versus deep-and-factorised, enabling assessment of augmentation effects across different inductive biases. The consistent direction of improvement across both networks strengthens confidence in the generalisability of the augmentation findings beyond any single architecture [18,20].

Seven geometric augmentation operations were evaluated: rotation at two orientations per case (90° and 270° for Case 1; 45° and 270° for Cases 2 and 3), horizontal flip, vertical flip, cropping, zooming, and 30° shearing. Applied to the abnormal classes only, combined basic augmentation increased dataset size by 77–129% across the three cases; Table 3 (§5.2) reports accuracy gains of approximately 4–24 percentage points for VGG-16 (most pronounced in Case 1, which had the most severe imbalance) and 5–6 percentage points for InceptionNet relative to unaugmented baselines.

Mass image augmentation employing basic techniques only helps in generating geometrically modified images, not entirely new and unique images. The analysis of individual augmentation technique contributions showed that not all techniques contribute positively: Rotation 270° and Shearing consistently result in better network performances across all cases; Rotation 90° was the best-performing first rotation for Case 1, while Rotation 45° performed best for Cases 2 and 3. The study further supported the hypothesis that combining the best performing strategies together results in better network performance than combining the worst performing strategies together.

Moreover, basic augmentation requires O(N) disk writes per technique, scaling storage and I/O linearly with the number of strategies applied — a practical overhead that grows as the augmentation pipeline expands.

The process of generating and saving images is automated, and GANs are capable of creating entirely novel images from genuine examples. The dataset was enhanced by 148%, 159%, and 139% for Cases 1, 2, and 3 respectively via WGAN synthesis (§5.4, Table 7). However, each batch of images generated with GANs has to be checked for quality before employing in the classification task.

While DCGAN was unable to learn and generalize for the given dataset, WGAN started to destabilize after approximately the 300th epoch; there exists a constraint on the amount of images that can be generated from GAN. Image quality screening is essential when employing GAN-generated data, as low-quality batches can degrade downstream classifier performance. The W-loss serves as a practical proxy for generation quality, with divergence between the score curves indicating training instability.

Depending on dataset size, basic augmentation techniques take ∼10–20 s, while GANs take around 1200 s (∼20 minutes) to generate one batch of output images.

The complementary weaknesses of each strategy motivated the hybrid approach: basic augmentation is fast but limited in variety and volume, while WGAN produces genuinely novel images but at high computational cost and with an epoch-based quality ceiling. The hybrid technique generated ∼11K images per case (∼35K in total across all three cases) and yielded ∼99% validation accuracy with both classifiers, constituting a practical and scalable augmentation pipeline applicable to medical imaging datasets of varying scale.

This study has several limitations. The evaluation is restricted to three disorder-pair case studies drawn from the VinDr-SpineXR dataset, and results may not generalise to other spinal pathologies or imaging modalities. The classifier set is limited to VGG-16 and InceptionNet; performance may differ with lightweight or transformer-based architectures. No external validation set was employed, so all results reflect held-out validation on the same dataset distribution. The epoch ceiling for WGAN image selection was determined by visual inspection rather than an automated stopping criterion, introducing a degree of subjectivity in the synthetic image selection process.

## 7. Conclusion

This paper presents an in-depth empirical comparison of augmentation techniques and their performance on the VGG-16 and InceptionNet deep learning architectures. Seven basic augmentation techniques (rotations at two orientations per case, horizontal flip, vertical flip, cropping, zooming, and 30° shearing) and deep learning augmentation using Generative Adversarial Networks were implemented. This study critically analyzed quality of generated images, time to generate images, contribution to network performance, and increase in dataset size to discern the best strategy. The study was repeated for each basic augmentation type to isolate the best performing basic techniques.

While each augmentation technique has merits and demerits, WGAN consistently outperformed combined basic augmentation, achieving VGG-16 validation accuracies of 97.07%, 95.09%, and 90.38% for Cases 1, 2, and 3 respectively, and InceptionNet validation accuracies of 91.85%, 93.19%, and 91.78% — an improvement of ∼9–27 percentage points over the unaugmented baseline.

Finally, the best performing basic augmentation techniques and WGAN were combined to form the hybrid augmentation strategy. This method combines suitable aspects of both data warping and oversampling, generating ∼11K images per case (∼35K in total) and achieving ∼99% validation accuracy with both VGG-16 and InceptionNet across all three case studies — the strongest result across all conditions evaluated. Limiting WGAN to its stable epoch range and supplementing with fast geometric transforms avoids the overhead of extensive GAN-only augmentation while maximising dataset size and classifier performance.

## 8. Future Work

Several directions merit investigation beyond the scope of this study. First, distribution-based metrics such as Fréchet Inception Distance (FID) and LPIPS should be applied to evaluate GAN-generated image quality more rigorously. Second, extending the methodology to additional radiological modalities, CT and MRI spine acquisitions, would broaden the applicability of the findings. Third, denoising diffusion probabilistic models and latent diffusion architectures represent a particularly compelling direction for future work: unlike GANs, diffusion models avoid mode collapse through their iterative denoising objective, and recent medical implementations, including RoentGen [16], have demonstrated high-fidelity, class-conditional synthesis for radiological images. A rigorous benchmarking of diffusion-based augmentation against the WGAN and hybrid pipelines evaluated here, using FID and LPIPS as quality measures, would constitute a timely and well-motivated extension of the present work.

## Data Availability

All data produced in the present study are available upon reasonable request to the authors

https://vindr.ai/datasets

